# Quantitative proteomics of coeliac gut during 14-day gluten challenge: low-level baseline inflammation despite clinical and histological normality predicts subsequent response

**DOI:** 10.1101/2020.05.04.20090977

**Authors:** Jorunn Stamnaes, Daniel Stray, Maria Stensland, Vikas K. Sarna, Tuula A. Nyman, Knut E. A. Lundin, Ludvig M. Sollid

**Author notes:** Corresponding authors: Jorunn Stamnaes, KG Jebsen Coeliac Disease Research Centre, Department of Immunology, Oslo University Hospital-Rikshospitalet and University of Oslo, 0424 Oslo, Norway., Ludvig M Sollid, KG Jebsen Coeliac Disease Research Centre, Department of Immunology, Oslo University Hospital-Rikshospitalet and University of Oslo, 0424 Oslo, Norway.

## Abstract

**Objective:** To shed light on gut mucosa processes provoked by gluten exposure in coeliac disease, we performed a quantitative proteomic analysis of duodenal tissue from well-treated coeliac patients before and after gluten challenge.

**Design:** We extracted and digested proteins from formalin-fixed paraffin-embedded tissue of 19 coeliac disease patients who had been challenged orally with gluten for 14 days. Protein identification and quantification was done by label-free quantitative mass spectrometry-based proteomics from total tissue and from laser capture microdissected epithelial cell layer. Proteomics data were compared with clinical, serological and histological data.

**Results:** At baseline, all patients were in clinical and mucosal remission (Marsh 0-1) except one (Marsh 3). After challenge, five patients reached Marsh 3 scores. Proteome analysis categorised these five and additionally two patients as responders. Already at baseline, responder patients differed from the remaining patients in their gut tissue protein composition with altered levels of inflammatory and enterocyte function proteins – the same proteins that changed upon gluten challenge. Patients classified as responders from the proteomic analysis also differed from the remaining patients at baseline, with mild crypt hyperplasia and a slight increase in blood inflammatory parameters and gluten specific CD4+ T-cell frequencies.

**Conclusion:** Despite clinical and histological remission, coeliac disease patients that develop a mucosal response after 14-day gluten challenge have already at baseline altered protein compositions of their gut tissue with signs of ongoing inflammation. Thus, apparently well-treated coeliac disease is frequently not fully quiescent with presence of low-grade anti-gluten immunity in gut mucosa.

## INTRODUCTION

Coeliac disease is a prevalent enteropathy caused by an aberrant, adaptive immune response towards dietary gluten proteins. Patients develop antibodies to deamidated gluten peptides and autoantibodies to transglutaminase 2 (TG2), the enzyme that catalyses deamidation. The disease is likely driven by activation the gut-homing gluten-specific CD4+ T cells that recognise deamidated gluten peptides in the context of the disease associated HLA-DQ2.5, DQ8 and DQ2.2 molecules.[1,2] These T cells are rapidly activated and undergo massive clonal expansion upon gluten ingestion. Dominant T-cell clones remain in the memory T-cell compartment of patients on a gluten free diet and are rapidly reactivated upon reintroduction of gluten.[3,4]

The small intestine is a dynamic tissue lined by a single layer of epithelial cells that is renewed every 3-5 days. New cells are generated from stem cells in proliferative crypts and differentiate to absorptive enterocytes or secretory cells such as goblet cells. Enterocytes migrate towards the villi tips and mature along the way, generating a zonal pattern of enterocyte function and phenotype.[5] The epithelial cell layer is surveyed by intraepithelial lymphocytes (IELs) and exposed to cytokines produced by IELs as well as lamina propria immune cells. The small intestinal lesion that develops in coeliac disease is characterized by increased IEL frequency, crypt hyperplasia, villous blunting and lamina propria plasma cell infiltration, and is considered to be a direct consequence of the adaptive immune response towards gluten.

For diagnosis, the lesion is usually graded according to a categorical classification score (Marsh score) based on IEL frequency, degree of crypt hyperplasia and villous blunting.[6] Continuous measures such as villus height to crypt depth ratio (Vh:Cd) and IEL counts are not routinely used for diagnosis, but are increasingly included as endpoints of clinical trials.[7] The only treatment available for coeliac disease is removal of gluten from the diet. Serum antibody levels to deamidated gluten peptides and TG2 drop within months [8,9], but normalisation of gut architecture can be slow and variable.[10,11] In a study of Norwegian adult coeliac disease patients, about 50% recovered with complete mucosal remission (Marsh 0-1) after one year of gluten free diet while close to 100% recovered within 8 years.[12] Reintroduction of dietary gluten (i.e. oral gluten challenge) was previously a mandatory part of the diagnostic work up in children, and is still required to diagnose subjects who have started a gluten free diet prior to examination. [13] While paediatric patients can be diagnosed with coeliac disease based on high serum of IgA anti-TG2 titers, evaluation of gluten-induced mucosal changes by Marsh score classification remains an essential part of the diagnostic work up in adult patients and paediatric patients with low levels of anti-TG2 antibodies.[13] Oral gluten challenge is also used in clinical trial settings where changes in Marsh score and Vh:Cd ratio serve as endpoints to evaluate drug efficacy.

The magnitude and kinetics of the mucosal response to oral gluten challenge can vary between and within studies. To understand what occurs in the intestinal tissue in response to gluten we should consider not only isolated immune cell populations, but also stromal compartments including epithelial cells which change dramatically upon tissue remodelling. A limited number of studies have performed global analysis of total coeliac biopsy material, as analysis is challenging due to data complexity and absence of cellular and spatial resolution.[14-17] Advances in mass spectrometry (MS)-based analysis of proteins extracted from FFPE biopsies is paving way for the use of archival clinical material collected for pathology assessment to perform tissue proteome analysis.[18] We recently demonstrated that proteome analysis of FFPE tissue sections from untreated and treated coeliac disease patients accurately captures disease hallmarks and processes that we know change upon disease remission.[19] By use of laser-capture microdissection (LCM), specific regions or compartments can be isolated from FFPE tissue sections for proteome analysis. Such an approach can provide spatial or cell-type resolved proteome information from miniscule amounts of FFPE tissue material.

Here we have analysed FFPE tissue from a well-characterised patient cohort of treated coeliac disease patients subjected to a 14-day gluten muesli-bar challenge.[20] All patients but one were in complete mucosal and clinical remission at baseline, but the mucosal response by day 14 varied. Our aim was to characterise processes that occur early in response to gluten to improve our understanding of the mucosal response to gluten challenge. We performed MS-based proteome analysis with label-free quantification of total tissue and LCM-isolated epithelial cell compartment and correlated our findings with available clinical and serological parameters. Our approach revealed presence of slight mucosal alterations already at baseline in patients that developed strong mucosal response by day 14. This explains why patients with seemingly similar clinical starting point responded differently to the same gluten challenge regime.

## MATERIALS AND METHODS

### Patient samples and clinical data

The 14-day oral gluten challenge and patient cohort has previously been described in detail.[20] In this study we have utilized tissue sections from FFPE biopsy blocks from 19 patients who completed the gluten challenge study. We compared our proteome data to previously published clinical data and clinical biochemistry measurements from blood samples collected at baseline before challenge. Gluten specific CD4+ T-cell frequencies in gut (baseline and day 14) and blood (baseline and day 6) for seven patients were previously reported. [3] (**Online supplementary Table 1**)

### Laser capture microdissection

Eight micrometre FFPE tissue sections were mounted on PEN-slides (Zeiss), air dried, deparaffinised with xylene and sequentially rehydrated in graded ethanol followed by brief staining with haematoxylin. Tissue regions were collected using a PALM MicroBeam laser capture microdissection system (Carl Zeiss MicroImaging, Munich, Germany) and samples were collected in 0.5ml adhesive caps (Zeiss).

### Protein digestion and mass spectrometry analysis

Fifteen 5μm FFPE sections were collected in a tube for total tissue protein extraction and digestion. Total tissue and LCM samples were dissolved 50mM ammonium bicarbonate and 0.1% ProteaseMAX Surfactant (Promega). Tissue samples were boiled and sonicated again to remove formalin crosslinks followed by protein disulphide bond reduction and cysteine alkylation prior to over-night digestion with trypsin. Tissue digests were separated on an Easy nLC1000 nano-LC system coupled to an Orbitrap mass spectrometer (ThermoElectron, Bremen, Germany) for data dependent acquisition (top 10 most intense peaks for MS/MS). (**Online supplementary Table 2**)

### Protein identification, quantification and data analysis

MS raw files were processed in the MaxQuant environment [21] (version 1.6.1.0) with the integrated Andromeda search engine [22] for peptide and protein identification. MS/MS spectra were searched against a UniProtKB FASTA database of the human proteome for protein identification. Label-free protein quantification was performed using the MaxLFQ algorithm also integrated in MaxQuant. [23] Further statistical and bioinformatical analysis was performed in Perseus [24] (version 1.6.2.2) and the R framework (https://www.r-project.org/). Mass spectrometry proteomics data have been deposited to the ProteomeXchange Consortium [25] via the PRIDE partner repository with the dataset identifier PXD(will be updated with accession number).

Further details can be found in the **online supplementary materials and methods**.

## RESULTS

### Classification of coeliac disease patients as responders and non-responders to gluten challenge based on their small intestinal proteomes

We collected tissue sections from FFPE biopsies used for histology assessment of 19 patients before and after a 14-day oral gluten challenge.[20] Shotgun, label-free quantitative proteome analysis of total tissue samples quantified 4301 proteins (**online supplementary figure 1A, B**). Unsupervised clustering segregated 8 biopsies along principal component 1 (PC1) of which 7 were collected from patients after challenge (**Figure 1A, online supplementary figure 1C B**). From this clustering we classified our patients as “responders” (n = 7) or “non-responders” (n = 12) to gluten challenge. Among the responders were all 5 patients classified with Marsh 3 lesion after challenge. In addition, two patients graded with Marsh 1 lesion after challenge were classified as responders based on proteome expression (**Figure 1B and online supplementary table 1**)

**Figure 1.**
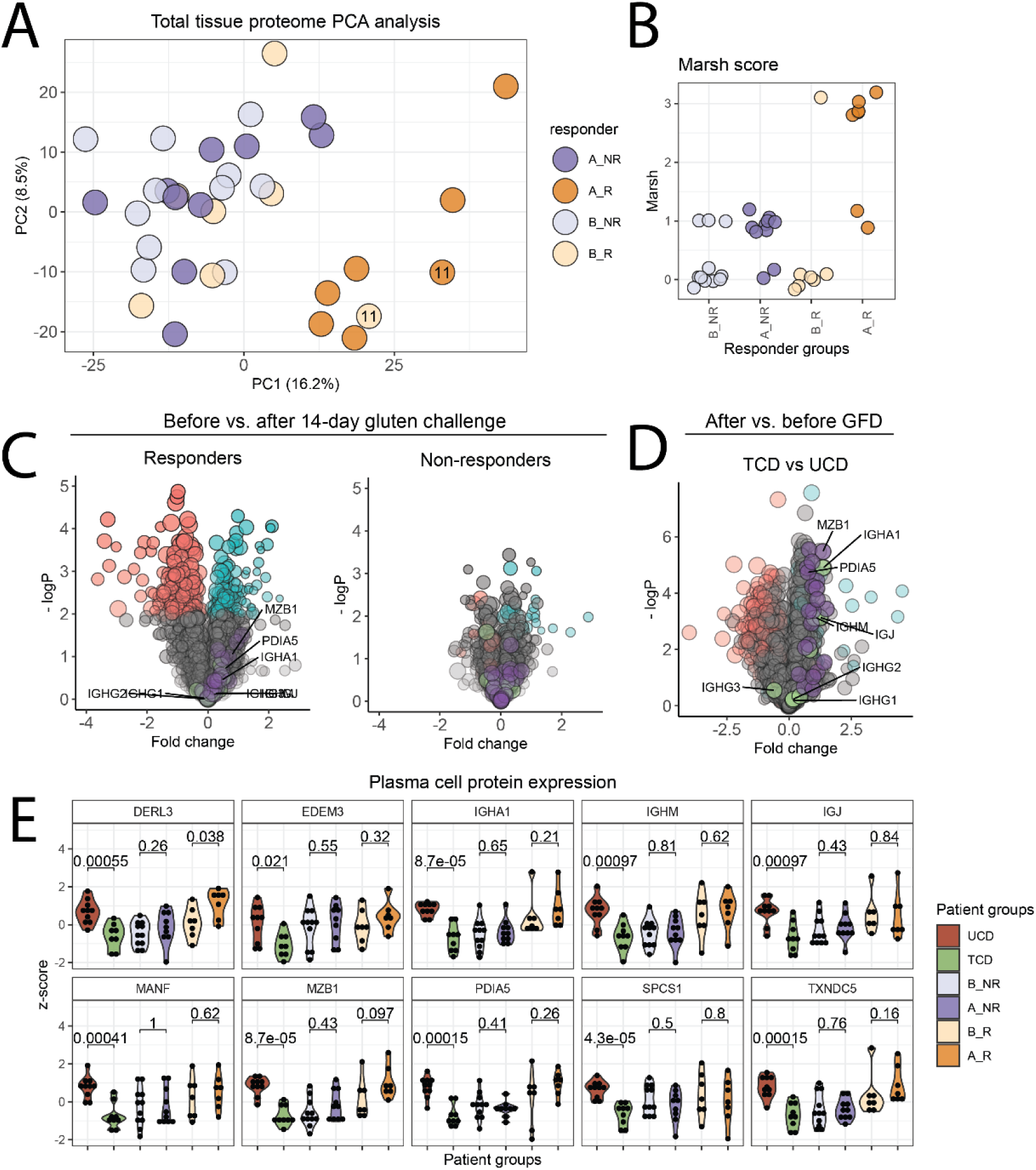
Stratification of coeliac disease patients to 14-day gluten challenge based on small intestinal proteome expression. **A**.PCA plot of total tissue samples show separation of patients along PC1 which accounts for 16.2% of the total variation. Responder after challenge patients (A_R) separate from the remaining patients. P11 = patient CD442 **B**. Marsh score for patients classified as responders and non-responders before and after challenge based on PCA separation in A. **C**. Volcano plot of fold change protein expression comparing responder samples and non-responder samples before and after challenge (Two-sample Student’s t-test, FDR = 0.05). **D**. Volcano plot of fold change for UCD and TCD samples from Tutturen et al.[19] **D and E** Proteins with significant change in responders after gluten (Two sample Student’s t-test p < 0.05) are indicated in blue (up, n = 115) and red (down, n = 143). Immunoglobulin genes are shown in green, other plasma cell derived proteins in purple. **E**. Comparison of z-scored expression of selected plasma cell derived proteins for the different patients groups (Two-sample Student’s t-test). UCD and TCD data are from [17]. B_R = before responders, B_NR = before non-responders; A_R = after responders; A_NR = after non-responders

Many of the proteins that drove separation of samples along PC1 are of neutrophil/ myeloid or mature enterocyte origin (**online supplementary figure 1C**). For responders we found differential expression of 258 proteins in response to gluten, but no significant changes for non-responders (**Figure 1C**). We have previously reported increased expression of neutrophil/myeloid derived proteins and decreased expression of mature enterocyte proteins in untreated coeliac disease (UCD) Marsh 3 compared to treated coeliac disease (TCD) Marsh 0-1.[19] A hallmark of UCD Marsh 3 is lamina propria plasma cell abundance, which is captured in tissue proteome datasets as immunoglobulins (**Figure 1D, green**) and as proteins involved in protein synthesis and ER stress (**Figure 1D, purple**). We observed only modest changes in expression of such proteins in response to gluten challenge for both responders and non-responders (**Figure 1C, E**). This observation suggests that plasma cell infiltration may follow a slower kinetic and is uncoupled from development of Marsh 3 lesion in response to gluten exposure.

### Epithelial proteome expression separates responders from non-responders after gluten challenge

To confirm the epithelial cell contribution in our total tissue dataset and address how the epithelial proteome responds to gluten exposure, we used LCM to specifically isolate and analyse the epithelial cell layer compartment (**Figure 2A**). To validate our approach, we first compared samples collected from three distinct tissue compartments; epithelial villi, epithelial crypts and lamina propria. These samples separated according to their anatomical origin, which confirms that we successfully captured and obtained informative datasets from small tissue regions (**online supplementary figure 2**). We next collected and analysed two cohorts of epithelial cell layer samples spanning the entire crypt-villus axis (**online supplementary materials and methods and online supplementary figure 3A**). In both datasets, samples separated clearly as responders or non-responders (**online supplementary figure 3B**). Combining the two datasets, we found clear separation of responder after challenge samples similar to our total tissue analysis (**Figure 2B, online supplementary figure 3C and E**). Samples from baseline biopsies of patient P11 (CD442, Marsh 3) were skewed towards but did not cluster with after challenge responder samples (**online supplementary Figure 3C)**. The clustering of baseline biopsies from P11 in our total tissue analysis therefor likely reflect contribution from lamina propria-derived proteins (**Figure 1A**).

**Figure 2.**
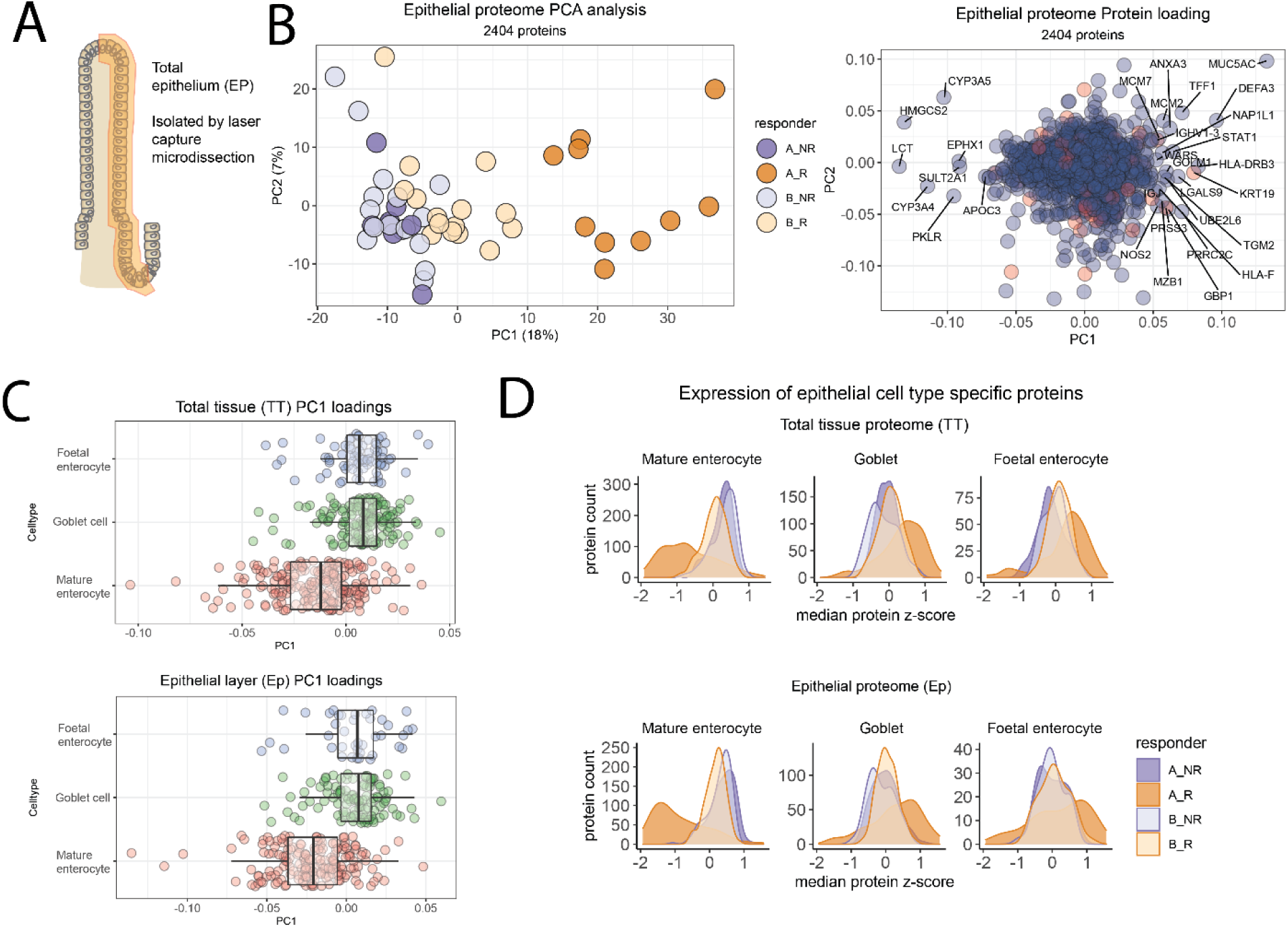
Analysis of total epithelial cell layer proteome from patient biopsies from before and after gluten challenge. **A**. Depiction of epithelial cell layer isolated by LCM **B**. PCA analysis based on expression of 2404 proteins separate samples from responder after challenge biopsies (A_R) from other samples. Right; protein loadings that drive the separation (blue, razor + unique peptides >1, red = razor + unique = 1) **C**. Distribution of proteins mapped to epithelial cell types along PC1 for total tissue and epithelial proteome data **D**. Expression of cell type specific proteins per responder group from total tissue (TT) or epithelial proteome data (Ep) (median z-score protein expression). B_R = before responders, B_NR = before non-responders; A_R = after responders; A_NR = after non-responders.

To estimate the epithelial cell layer composition and phenotype in our biopsies we mapped our proteome data to cell-type specific gene sets derived from published single cell RNA-sequencing analysis of small intestinal epithelial cells, choosing datasets for “mature enterocytes”, “foetal-like enterocytes” and “goblet cells” (**online supplementary table 3**).[26,27] The distribution of cell-type specific proteins along principle component 1 (PC1) show that mature enterocyte proteins are skewed towards non-responder samples while goblet cell and foetal enterocyte proteins distribute towards after challenge responder samples (**Figure 2C**), in agreement with the reduced expression of mature enterocyte proteins and increased expression of foetal enterocyte and goblet cell proteins in responder after challenge samples (**Figure 2D and Supplementary Figure 3D**). Our data show that changes in epithelial proteome expression alone can classify our patients as responders or non-responders to gluten challenge.

### Patients that respond strongly to gluten challenge differ from non-responders at baseline

In the epithelial proteome dataset, we found differential expression of 658 proteins after gluten challenge for responder samples, and no clear changes for non-responders (**Figure 3A, panel 1 and 2**). However, comparison of baseline samples for responders vs. non-responders revealed differential expression of many or the same proteins that changed in responders after challenge (**Figure 3A, panel 3**). The fold change expression for differentially expressed proteins before vs. after challenge for responders correlated with the fold change expression for the same proteins when comparing responders vs. non-responders at baseline (R = 0.49, p = 2.2E-16) (**Figure 3B panel 1**). No correlation was seen with before vs. after challenge for non-responders (**Figure 3B panel 2**).

**Figure 3.**
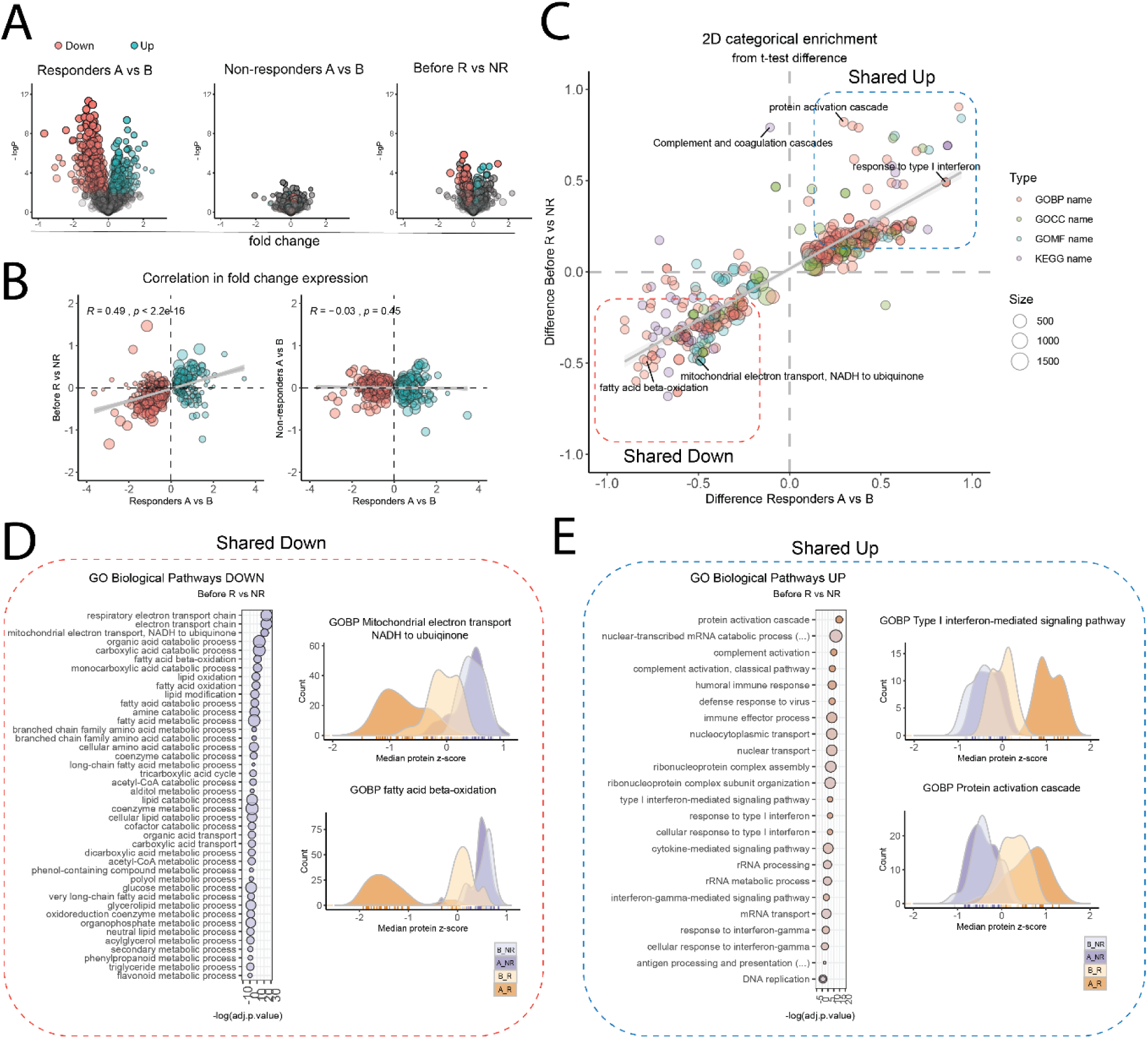
Differential protein expression in responders and non-responder samples at baseline resemble changes that occur in response to gluten challenge. **A**.Volcano plot of fold change protein expression comparing responder samples and non-responder samples before and after challenge, and baseline responder vs. non-responder samples. Proteins with p< 0.05 (responder samples, before vs. after challenge, student’s two-sample t-test, FDR <0.05) are indicated in blue (up, n = 324) and red (down, n = 334) in all three volcano plots. **B**. Correlation in fold change protein expression for after vs before challenge responder samples (A_R vs B_R, Pearson correlation) and baseline responders vs. non-responder samples (B_R vs. B_NR, Pearson correlation). **C**. 2D enrichment analysis for biological processes based on fold change protein expression. Color indicate type of pathway, size indicate number of genes in the pathway. **D and E**. Comparison of expression for GOBP among Shared Down pathways (**D**) and Shared Up pathways (**E**). Expression of proteins mapped to GOBP pathways was compared between responder groups (ANOVA and Tukey’s honest significance test).Pathways are ranked according to adjusted p-value comparing responder vs non-responders. Size indicate number of proteins in the pathway. Protein count distribution based on protein expression (median z-score) is compared for responder groups for selected pathways. (B_R = before responders, B_NR = before non-responders; A_R = after responders; A_NR = after non-responders)

To address what biological pathways or processes the differentially expressed proteins represent, we performed 2D enrichment analysis for biological pathways (**online supplementary materials and methods**).[28] Enriched pathways correlated for responders before vs. after challenge and responders vs. non-responders before challenge (**Figure 3C**).

Many of the pathways that were diminished in responders after challenge (Shared Down) represent mature enterocyte processes. Proteins from pathways that involve fatty acid metabolism and mitochondrial function were most differentially expressed between responders vs. non-responders at baseline (**Figure 3D**). Proteins mapped to “GOBP Mitochondrial electron transport, NADH to ubiquinone” and to “GOBP fatty acid beta-oxidation” showed lowest protein expression for responders after challenge but with clear difference between responders and non-responders at baseline (**Figure 3D**). These differences did not depend on the presence of samples from patient P11 (Marsh 3 before challenge) (**online supplementary figure 4**). Thus, enterocyte function was altered already prior to gluten challenge in responders.

Among pathways that were increased in responders after challenge (Shared Up), innate immune activation, complement and interferon/cytokine responses were increased in responders compared to non-responders at baseline (**Figure 3E**). Proteins that mapped to “GOBP Cellular response to type-I interferon” showed higher expression in responders compared to non-responders at baseline but increased strongly in responders after challenge. Proteins that mapped to “GOBP protein activation cascade” had higher expression in responders than non-responders before challenge but here the increase after challenge was only modest. In summary, our proteome analysis of the epithelial cell compartment revealed that responders differ from non-responders already at baseline before challenge.

### Villus height crypt depth ratio correlates with mature enterocyte protein expression and differs for responders and non-responders at baseline

The similar distribution of epithelial cell-type specific proteins in our epithelial and total tissue data signified that our LCM isolated epithelium faithfully represent total biopsy epithelium (**Figure 2B and online supplementary figure 5**). Skewed cell type protein expression in baseline responder biopsies may therefore reflect small changes in cell type or phenotype ratios, or global changes along the entire crypt-villus axis (**Figure 4A**). Marsh score is a categorical classification that only captures gross morphological changes and IEL frequency. We therefore compared Vh:Cd ratio with cell-type protein expression per patient (**online supplementary table 4**). We observed positive correlation with mature enterocyte protein expression and negative correlation with foetal enterocyte and goblet cell protein expression (**Figure 4B**). Vh:Cd ratio was significantly lower in responder compared to non-responder biopsies at baseline (**Figure 4C**). Disregarding patient P11 (Marsh 3 at baseline), we see that 4 of 12 non-responder biopsies were Marsh 1 at baseline compared to none of the responder biopsies. This agrees with higher baseline CD3+ IEL frequency in non-responders (**Figure 4C**). Responder biopsies may therefore have mild crypt hyperplasia at baseline but this is independent of CD3+ IEL number.

**Figure 4.**
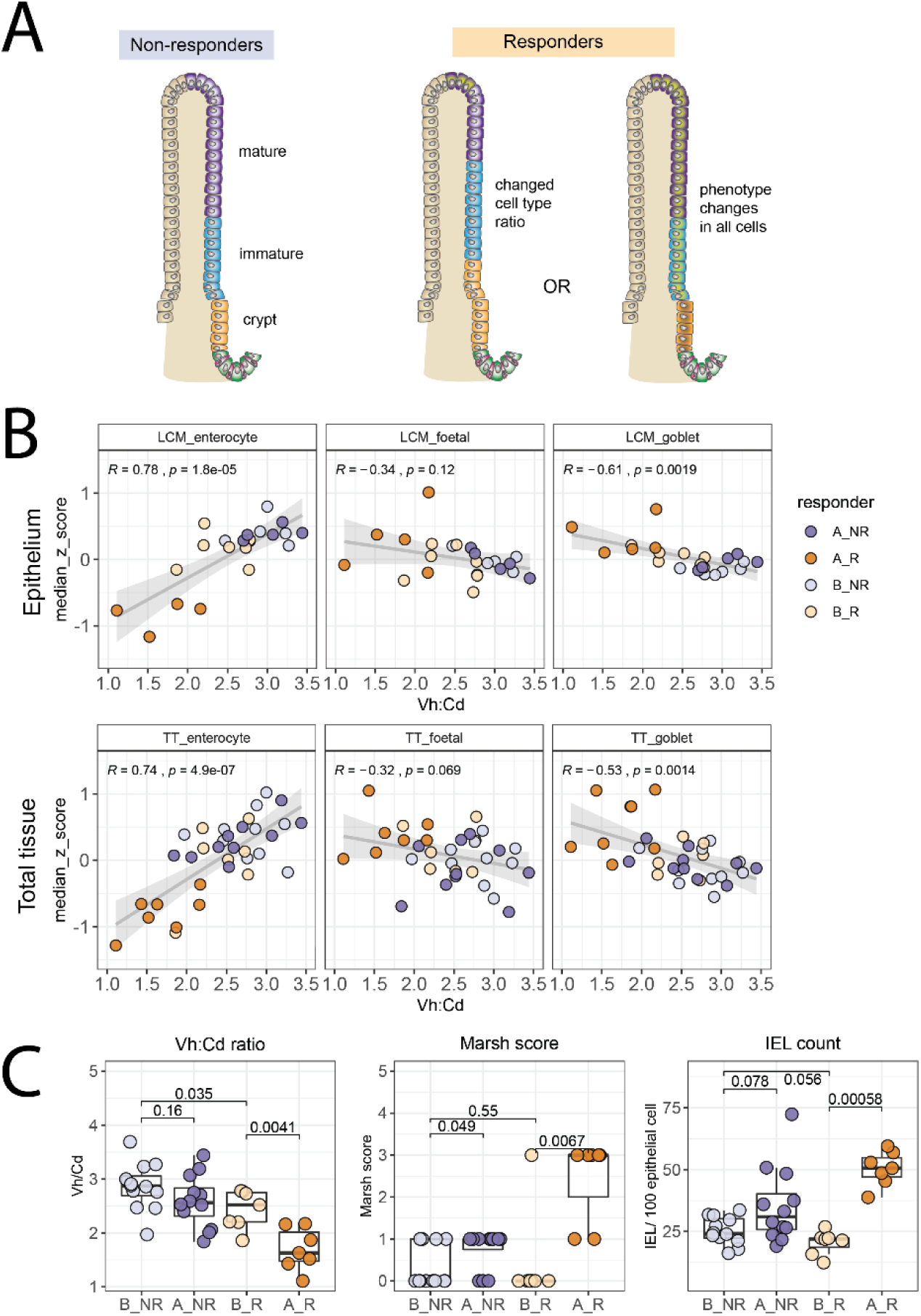
Villus height to crypt depth ratio correlates with cell type protein expression and differs for responders and non-responders at baseline. **A**. Depiction of scenarios that can explain altered cell type protein expression in responder patients at baseline **B**. Correlation between Vh:Cd ratio and epithelial cell type protein expression per patient (median z-scored expression) from epithelial (LCM) and total tissue proteome data (TT). **C**. Comparison of Vh:Cd, Marsh score and CD3+ IEL count for responder groups (Wilcoxon ranked test)

To remove an effect of crypt compartment changes, we carefully isolated the top 50% epithelial layer by LCM from a subset of responder and non-responder baseline biopsies (apical epithelial layer, **Figure 5A**). We isolated a smaller tissue areal per sample compared to total epithelial samples, resulting in a lower number of quantified proteins in this dataset (**online supplementary figure 6**). Unsupervised clustering (PCA) separated responders from non-responders (**Figure 5B and online supplementary figure 6**). However, here we did not see reduced expression of mature enterocyte proteins in responder samples as observed in the total tissue and epithelial proteome datasets (**Figure 5C**). This supports the notion that reduced expression of mature enterocyte proteins at baseline in responders is a result of mild crypt hyperplasia.

**Figure 5.**
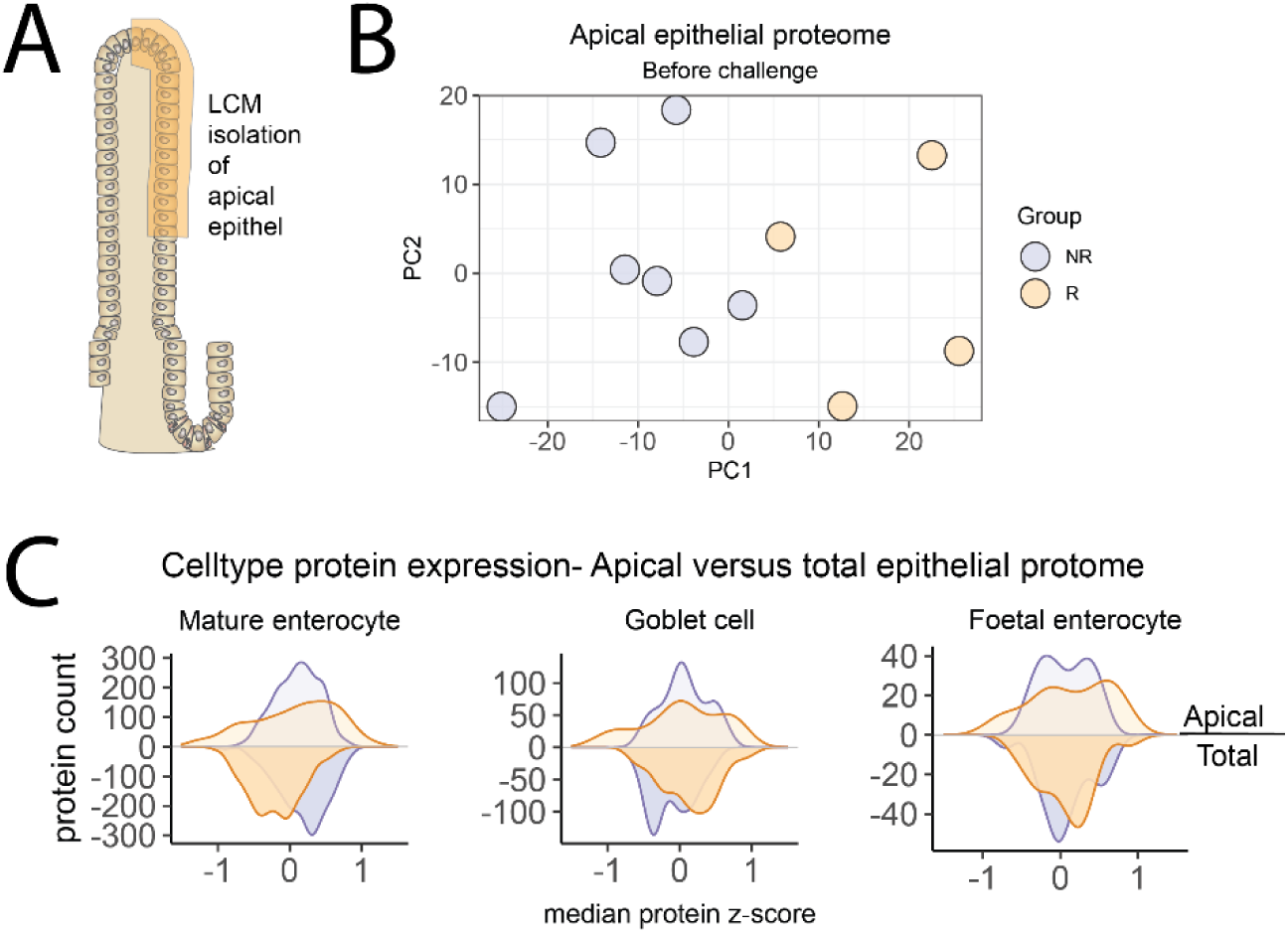
Mature enterocyte protein expression is not skewed in the apical epithelial proteome of responders. **A**. Depiction of apical epithelial cell layer isolated by LCM from biopsies collected at baseline before challenge **B**. PCA analysis based on the expression of 1151 proteins show that baseline biopsies from responders separate from non-responders **C**. Distribution of cell type specific protein expression in apical epithelial proteomes compared to total epithelial proteome. For this comparison, total epithelial proteome data were z-scored across baseline samples only, and expression values for the same proteins that were mapped to the apical proteome dataset are shown.

### Presence of low level inflammation and immune activation in responders at baseline

Pathways indicative of acute phase response or complement activation were enriched in the epithelial proteome of responders at baseline. Proteins that map to the KEGG complement and coagulation cascade pathway had higher expression in responder samples both in our epithelial proteome (**Figure 6A, B**) and apical epithelial proteome datasets (**Figure 6, D**). Some of these proteins, such as complement and fibrinogen components, are abundant in serum. However, we observed no difference in haemoglobin protein abundance between responders and non-responder samples, which would argue against increased blood vessel contamination in responder samples as the reason for this observation (**Online supplementary figure 7**). We next addressed whether other available variables differed for responders and non-responders prior to gluten challenge. We observed a slight increase in serum ALAT, ALP, ASAT, Crp, Ferritin, GT, Hb and plasma TNF-α in responders while serum transferrin was decreased (**Figure 6D, online supplementary table 5 and online supplementary figure 8**). This pattern of clinical biochemistry parameters supports the notion of low-level inflammation at baseline in responders. The values of these biochemistry parameters are however within the range of normal values and can therefore not alone be used to predict responders.

**Figure 6.**
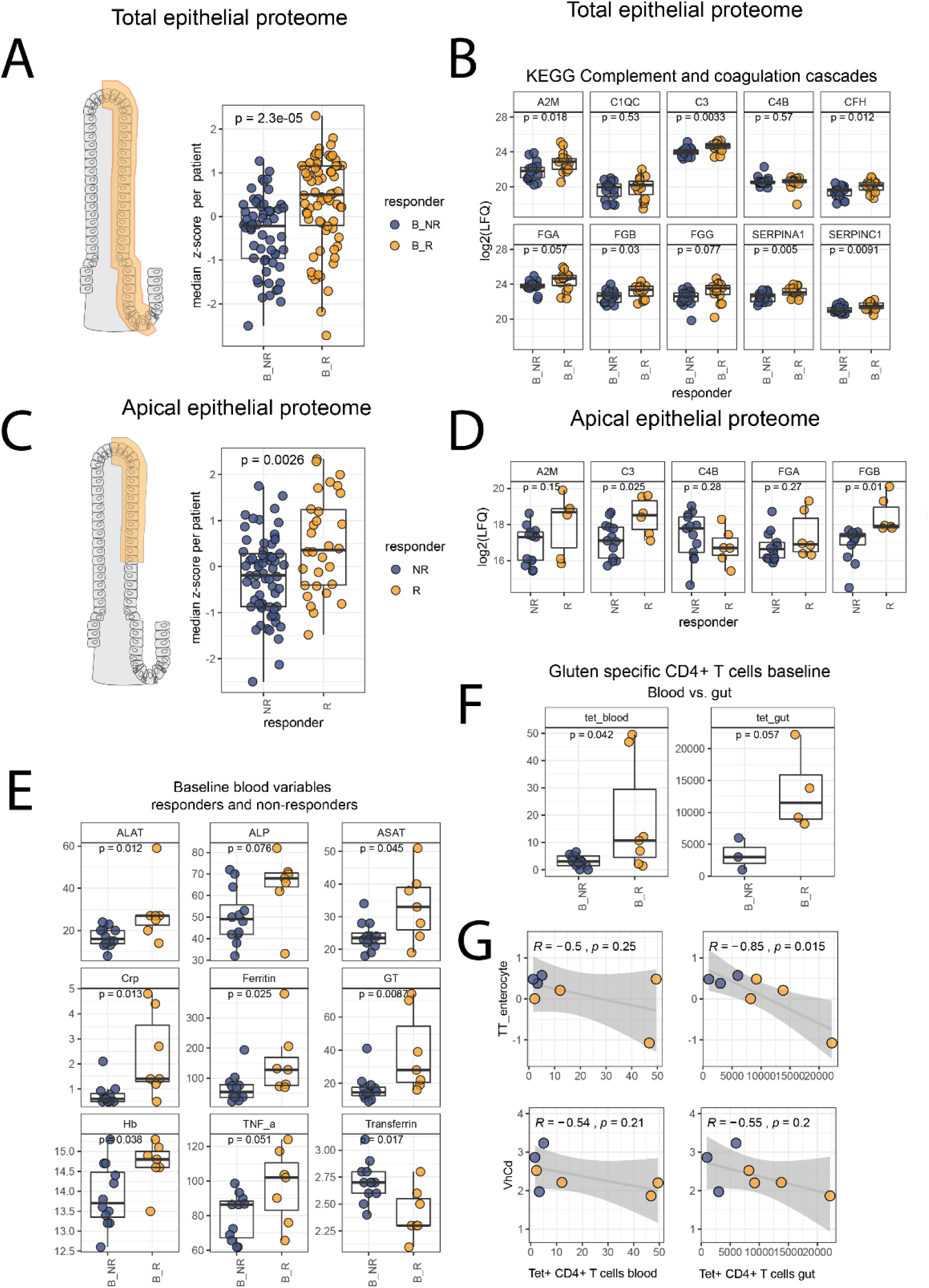
Presence of low-level epithelial proteome and serological markers of inflammation at baseline in responders agree with presence of low level gluten immunity at baseline. **A, C**. Comparing expression (z-score) of proteins that map to KEGG complement pathway in total epithelial proteome (A) and apical epithelial proteome (C) for responder versus non-responder patients at baseline (Welch t-test). **B, D**. Comparison of log2(LFQ) expression values for individual proteins (Welch t-test) from total epithelial proteome samples (B) and apical epithelial proteome samples **E**. Comparison of clinical biochemistry and cytokine response measured a baseline before challenge for responders and non-responders (Wilcoxon ranked test). **F**. Comparison of frequency of gluten-specific gut homing effector memory T-cells per million CD4+ T-cells in blood at baseline and gluten specific CD4+ T-cells per million CD4+ T-cells from gut biopsies collected at baseline. Data for patients P2, P5, P7, P11, P12, P13, P14, P15 is shown (Wilcoxon ranked test) **G**. Correlation between baseline median mature enterocyte protein expression (from total tissue proteome dataset) and Vh:Cd ratio with gluten-specific CD4+ T-cell frequencies in gut and blood (Pearson’s correlation)

Twelve of 15 analysed patients had >2 fold-change of gluten specific gut-homing effector memory CD4+ T cells in blood by day 6 compared to baseline.[20] We compared Vh:Cd ratio and mature enterocyte protein expression data with numbers of gluten specific CD4+ T cells in blood and gut biopsies at baseline (**Online supplementary table 1**). T-cell numbers from gut biopsies were available for seven of the patients from this cohort and comparison was made for these patients only (**Figure 6F**). Frequency of gluten specific T cells in blood correlated both with Vh:Cd ratio and mature enterocyte protein expression at baseline (**Figure 6G**) and day 6 although this correlation was mainly driven by high T-cell numbers in a small number of patients (**Online supplementary figure 9**). Vh:Cd ratio and mature enterocyte protein expression also correlated with the number of gluten specific T-cells in gut at baseline (**Figure 6G**) and at day 14 after challenge (**Online supplementary figure 9)**. Although the analysis is based on a limited number of patients, this suggests that responder patients with ongoing low-level inflammation at baseline may have a higher number of gluten-specific CD4+ T cells, and particularly so in the gut compartment.

## DISCUSSION

To characterise the mucosal recall response to gluten in coeliac disease, we performed a proteomics analysis of FFPE biopsy tissue collected from well-treated coeliac patients before and after a 14-day gluten challenge.[20] Based on tissue proteome expression we classified seven of 19 patients as responders to gluten, which included the five patients classified with Marsh 3 lesion, in addition to two patients with Marsh 1 lesion. This discrepancy likely relates to the inaccuracy of Marsh classification for biopsies with minor alterations and argues that our approach more robustly groups similar patients together. Our proteomics analysis revealed epithelial inflammation and changes indicative of mild crypt hyperplasia in responders at baseline. These findings were confirmed by slight alterations in Vh:Cd ratio and serum profile of responders. Our data support the notion that baseline differences explains the variation in mucosal response within gluten challenge cohorts. While these differences may go unnoticed upon conventional clinical evaluation, they are likely relevant for gluten challenge outcome as assessed by changes in Marsh score.

Both gluten dose and challenge length influence mucosal response kinetics to gluten. Two comparable 14-day gluten challenges yielded very different outcomes in terms of Marsh score; Leffler et al reported Marsh 3 for 13/19 patients compared to 5/19 patients in the study cohort we have analysed.[20,29] Such variation poses a challenge in particular for clinical trials when changes in mucosal histology serve as an endpoint to evaluate drug efficacy. Long challenge regimes are therefore required to ensure that a large fraction of patients indeed develop intestinal pathology. Use of continuous measures such as IEL count and Vh:Cd ratio to assess histological remission and baseline may increase the resolution and improve evaluation of mucosal response to challenge. While our proteomics data showed strong agreement with Vh:Cd ratio, the IEL count at baseline was lower for responders compared to non-responders. CD3+ IEL count at baseline was therefore not a positive predictor of mucosal response to gluten challenge in the cohort we have studied. Notably, CD3+ IEL count reveal no information about the composition of the IEL compartment, which may be of relevance for the kinetics or magnitude of the mucosal response to gluten.

LCM isolation and analysis of the epithelial cell compartment confirmed that the epithelial proteome composition alone reveals differences between patients at baseline before gluten challenge. Mature enterocytes proteins have previously been shown to be reduced in UCD biopsies and tissue APOA4 and Ki-67 mRNA expression ratio has been proposed as a molecular measure of Vh:Cd ratio before and after 6-week gluten challenge.[30] Reduced expression of mature enterocyte proteins in our dataset together with reduced Vh:Cd ratio indicates that responders have mild crypt hyperplasia at baseline. These epithelial compartment differences may be too subtle for reliable detection by methods that monitor expression of a few proteins such as immunohistochemistry. As proteome analysis considers expression of thousands of proteins, we can assess the sum of small difference for sets of proteins. We chose to map our proteome data to gene sets that represent mature enterocytes, foetal enterocytes and goblet cells. These proteins are highly likely to be of epithelial cell origin in contrast to proteins that map to proliferative cell types such as transit amplifying cells, where signal in our dataset also may come from IELs that proliferate in response to gluten.[31] We interpret the increased expression of foetal enterocyte and goblet cell specific proteins in responders after challenge as a proxy for epithelial inflammation. In a mouse model of small intestinal parasitic crypt infection, CD4+ T cell derived interferon-gamma resulted in reduced expression of mature enterocyte mRNA and increased expression of foetal enterocyte and goblet cell mRNA in the small intestinal epithelium. [26] We have not investigated whether the slight increase in goblet cell protein expression reflects measurable numeric change in goblet cell frequency but consider it here as a measure of tissue state. The good agreement between our total tissue and LCM isolated datasets strengthens our confidence in the validity of the differences we observe.

The increase in complement pathway proteins in the epithelium and inflammatory serum biochemistry supports the notion of low-level inflammation at baseline in responders. Enterocytes can express complement and acute phase proteins in response to damage or inflammatory stimuli.[32,33]. As we measured proteins and not RNA transcripts, we can however not exclude that complement proteins in the epithelium may derive from serum deposition, as previously demonstrated in UCD mucosa.[34] Conceivably, the mucosal pathology of coeliac disease results from downstream consequences of a CD4+ T-cell response to gluten. These T cells respond extremely rapidly to gluten as the T-cell derived cytokine IL-2 can be measured in blood within hours of gluten consumption.[35] Gut-homing gluten specific CD4+ T-cells peak in blood on day 6 after gluten challenge. While 12 of 15 analysed patients from the cohort we here have studied were considered as “responders” based on fold change in blood T-cell frequency, only 5 patients were mucosal responders based on Marsh score (of which one were Marsh 3 already at baseline). Fold change in tetramer positive cells gives only a relative measure of change, and it is likely that for a mucosal response, the baseline number of T cells is more critical. Indeed, albeit from a small number of patients, we see a trend towards higher baseline numbers of gluten specific T cells in our tissue proteome responders compared to non-responders. The proteome differences that separated mucosal tissue of responders and non-responders at baseline was qualitatively similar to the changes induced by gluten challenge in responders. These observations suggest that some patients which appear to be in complete remission, may still have ongoing low-grade anti-gluten immunity in the gut mucosa.

In summary, we here demonstrate that MS-based proteomics analysis of FFPE intestinal tissue provides important information that is not evident from regular histology assessment. The use of laser capture microdissection to obtain spatially resolved proteome information opens the possibility to study tissue sub-compartments, as we here have shown for the epithelial cell layer. We obtained meaningful information from very small amounts of tissue even with relatively limited proteome depth. This demonstrates the strength of addressing a specific question using a well-defined patient cohort. The field of MS-based analysis of clinical FFPE material is developing rapidly and currently receiving much attention in particular in the field of cancer diagnostics.[18] While the applicability for diagnosis may yet be limited, this approach represent a highly valuable complementary tool to classical tissue histology analysis.

## Data Availability

Proteomics raw data and affiliated data are available in proteomeXchange repository as reported in the materials and methods section. All data is available from the authors upon request.

## ACKNOWLEDGEMENTS

We thank Kjersti T Hagen for collection of FFPE tissue sections and Eivind Gard Lund for help with initial data analysis. The PALM Microbeam is part of the Advanced Imaging Core Facility and proteomics analysis was performed at the Proteomics Core Facility, both supported by the Core Facilities program of the South-Eastern Norway Regional Health Authority.

## FOOTNOTES

### Contributors

Conception and design: JS, VKS and LMS. Performed experiments and acquired data: JS, DS, MS. Data analysis: JS, DS. Interpretation of data: JS, DS, LMS. Provided patient material and clinical data: VKS, KEAL. Writing of manuscript: JS, LMS. Critical review of data and manuscript: all authors.

### Competing interests

KEAL has privately or via his employer been consultant during the two last years for Chugai Pharmaceutical, Bioniz Therapeutics, Immusant Therapeutics, Interexon Actobiotics, Amyra Biotech AG and Dr. Falk Pharma GMBH.

LMS has privately or via his employer been consultant during the two last years for Amagma Therapeutics, Chugai Pharmaceutical, Bioniz Therapeutics, Immusant Therapeutics, Interexon Actobiotics, UCB Biopharma, Merck and GSK. The other authors declare no conflicts of interest.

**Ethical approval** Patient material used in this study was collected as part of the clinical trial NCT02464150 for which ethical approval and patient consent were collected and previously reported [ref 20].

**Data availability statement** Proteomics raw data and affiliated data are available in proteomeXchange repository as reported in the materials and methods section. All data is available from the authors upon request.

